# Emotional intelligence and the perception of good leadership in healthcare: a mixed-methods study

**DOI:** 10.64898/2026.08.23.26361157

**Authors:** John W. Wilson, Angela Michaelis, Mary-Ellen Miller

## Abstract

**Objective:** To identify and rank the leadership traits most valued by medical staff in public hospitals, and to compare them with an established generic instrument and a generative artificial intelligence (AI) source.

**Design:** Sequential exploratory qualitative-quantitative (QUAL-QUAN) mixed-methods study: focus groups followed by an online ranking survey, with cross-comparison against the Northouse Leadership Traits Questionnaire (LTQ) and a ChatGPT-derived list (LAIT).

**Setting:** Major public hospital affiliated with Monash University, Melbourne, Australia, in 2023.

**Participants:** Twenty-four senior medical staff (16 men, 8 women; 18 clinicians, 6 administrators) recruited through opportunistic sampling.

**Main outcome measures:** Weighted ranking of the ten most desired leadership traits (Leadership Enabling Traits Survey, LETS); internal consistency (Cronbach’s α); agreement between LETS and LTQ self-scores; and strong overlap with the AI-derived list.

**Results:** The first most-weighted LETS traits were integrity (1.526), communication (1.435), compelling vision (1.404), emotional intelligence (1.040) and empathy (0.969) – the same five identified by the AI source. Integrity was weighted 3.6 times more heavily than rebelliousness (0.424). Both LETS and LTQ achieved Cronbach’s α >0.7. Unweighted total self-scores did not differ between LETS (77.4±7.6) and LTQ (78.4±6.9); weighted emotional intelligence related and other-trait sub-scores diverged significantly (p<0.001). Survey power was 45% at α=0.05.

**Summary Box:** *What this paper adds:* What is already known on this topic

- Emotional intelligence (EI) is consistently identified as an important attribute of effective healthcare leaders, and a range of generic leadership instruments are available (e.g. Northouse’s LTQ).
- Leadership-selection criteria in healthcare have historically been derived top-down from organisational, regulatory or academic frameworks rather than from the views of the clinicians who must follow those leaders.
- Existing instruments treat candidate leadership traits as equally weighted, even though clinical leaders intuitively rank some attributes (e.g. integrity) far above others. What this study adds

- 10-item, follower-derived, weighted Leadership Enabling Traits Survey (LETS) instrument was developed from focus groups with senior medical staff and ranked by survey, providing an empirically-weighted alternative to existing equally-weighted tools.
- top five LETS traits (integrity, communication, vision, emotional intelligence, empathy) matched the unprompted top five traits generated by a contemporary large-language model (LAIT), suggesting that EI-related attributes are a stable feature of leadership representations in both human and machine-generated sources.
- empirically-derived weights revealed material divergence between context-specific (LETS) and generic (LTQ) instruments when EI-related and non-EI traits were considered separately, supporting the case for situation-specific leadership tools rather than universal scoring.

## Introduction

There is currently a global crisis in healthcare, associated with escalating costs, manpower shortage and increasing population longevity. The NHS has seen a sequence of inquiries that have highlighted the need for capable leaders to develop and enact appropriate policies to ensure optimal health outcomes with cultural change (1–3).

The COVID-19 pandemic presented a major and unexpected load-challenge for all healthcare systems. The unprecedented agility shown demonstrated potential for good decision-making by healthcare leaders in a crisis, however, post pandemic, healthcare culture has not improved, having been described as based on *‘disciplinary action, bullying, whistleblowing and recruitment and career progression’* (3–5). Analyses of pandemic leadership argue that effective crisis response depended less on individual command than on leaders’ capacity to build and mobilise a shared group identity, fostering a collective sense of “we” rather than directing from a position of “I”(6). It has been asserted that leaders themselves are responsible for generating and maintaining an optimal organisational culture (7).

The findings also speak to the selection of leaders who are responsible for good decision-making and organizational culture, which may benefit from better understanding of what healthcare leaders should look like (8). Among the qualities repeatedly linked to effective leadership is emotional intelligence (EI), popularised as a set of workplace competencies distinct from cognitive ability and of growing interest in healthcare (9).

The needs of followers are often not considered(10) when determining KPIs for appointed leaders. This is despite the fact that organisations routinely use ‘market research’ to understand the needs of their customers, and despite longstanding calls within the leadership literature to treat followers as active co-producers of leadership rather than passive recipients. From this perspective, followers’ experiences and evaluations are central to how effective leadership is defined(11). We hypothesize that senior medical staff can list ideal traits of leaders and rank them based on perceived importance.

This study aims to: i) use a mixed-methods research (MMR) approach to determine qualities (traits) of good leaders and rank them according to perceived importance; and ii) characterise prioritised traits by comparison with a) a published tool and b) an artificial intelligence source.

## Methods

### Study design

The MMR approach uses a sequential, exploratory qualitative-quantitative design with deductive drive (12, 13). The qualitative component identified subjective opinions about leadership descriptors (qualities, traits, behaviours) and then using an online survey, ranked them to assign a weight according to perceived importance as an assessment tool (LETS, Leadership Enabling Traits Survey). Participants self-assessed using LETS and a published, generic, leadership assessment tool (LTQ, Leadership Traits Questionnaire) (14).

**Setting and participants.** A cohort of medical staff from major public hospitals in Melbourne, Australia was invited to participate. The study was approved by the Monash University Human Ethics Research Committee (Project 35811, approved July 2023) and was supervised at Kings Business School, London. Subjects were recruited by opportunistic sampling from amongst Monash University-affiliated Senior Medical Staff. Of 24 subjects: 16 were male and 8 were female; 6 were primarily administrators who were not in clinical practice. All subjects approached accepted the invitation to participate. The sample was designed as an exploratory pilot, providing the evidence base for the quantitative survey. Survey data collection was limited to participants in focus groups to enable initial characterisation of traits (data linkage) (See supplementary material).

**Qualitative focus groups.** The method used is as previously described (15). The segmental, protocol nature of discussion and MMR integration is described below. After obtaining informed consent, investigator-initiated discussion with groups (each n ≤ 5 subjects) took place on the topic of leadership traits in healthcare.

Following discussion, subjects were asked to list their most / least desired leadership traits. Responses were then transcribed freehand into WORD_™_ in digital format then converted into a spreadsheet for entry into NVIVO_™_ (16). Thematic analysis was then undertaken to identify consistent streams (17).

**Quantitative survey.** The quantitative survey component used a closed-ended approach to rank the traits found in focus groups (18). The same 24 subjects were sent survey questions using a link to an anonymised data collector (Survey Monkey_™_) (19). Of the 24 subjects, *n*=19 responded to an email with a survey link. Participants then rated themselves using both LETS and LTQ.

**AI Comparator.** To obtain a Leadership scale of AI traits (LAIT), OpenAI’s ChatGPT (GPT-3.5) was interrogated on a single occasion on 22^nd^ July 2023(20). It was asked verbatim, to list the 10 best leadership traits and then to assign an importance weighting to each; no system prompt, plugins or fine-tuning were used, and inter-run variability was not assessed. The AI was used solely as a point-in-time comparator data source, and not for study design, analysis, or manuscript preparation. Outputs are reproduced verbatim in Supplement 1. (As a non-deterministic, version-dependent model, the LAIT list represents a single 2023 snapshot and is un-validated)

**Statistical Analysis**. The distribution of LETS and LTQ self-scores were tested for normality using the Kolmogorov-Smirnov and Shapiro-Wilk tests. Internal consistency for each instrument was assessed with Cronbach’s α, with α >0.70 taken as acceptable. Total and subscale self-scores (unweighted and importance-weighted) were compared between LETS and LTQ using linear regression; the coefficient of determination (R²) is reported for the weighted-versus-unweighted subscale comparison. Mean scores for LETS and LTQ were compared using a paired t-test (GraphPad Prism, version 11.0.2). Statistical significance was set at p<0.05. The survey sample was opportunistic and powered as an exploratory pilot; post-hoc power was 45% at α=0.05, and findings are therefore treated as hypothesis-generating.

## Results

### Identification of traits

It was apparent from focus groups that subjects treated EI as a separate trait to its defined components (Table 1). Preferred traits became more popular as the session progressed (top 10 were 51% of total preferred traits on completion versus 36% of total at beginning). Focus groups therefore enabled ‘coning down’ of concepts and preferred traits. The 10 traits were then ranked in the online survey (Table 1) by *n*=19 respondents of the 24 participants in focus groups.

**Table 1.** Ranked leadership traits and importance of weights from the follower-derived survey (LETS), the AI-derived list (LAIT) and the Northouse Leadership Traits Questionnaire (LTQ)

|  | **LETS trait** | **weight** | **Chat GPT (LAIT)** | **weight** | **LTQ** | **weight** |
| --- | --- | --- | --- | --- | --- | --- |
| 1 | Acts with integrity | 1.526 | Integrity | 1.071 | Articulate: Communicates effectively with others | 1.196 |
| 2 | Excellent communication skills | 1.435 | Communicator | 1.071 | Trustworthy: Is authentic and inspires confidence | 1.127 |
| 3 | Clearly articulates a compelling vision | 1.404 | Visionary | 1.071 | Perceptive: Is discerning and insightful | 1.027 |
| 4 | Has good emotional intelligence | 1.040 | EI | 1.071 | Dependable: Is consistent and reliable | 0.878 |
| 5 | Shows empathy | 0.969 | Empathy | 1.071 | Persistent: Stays fixed on the goals, despite interference | 0.820 |
| 6 | Is a critical thinker | 0.960 | Problem solver | 0.952 | Self-confident: Believes in himself/herself and his/her ability | 0.768 |
| 7 | Is a teambuilder | 0.949 | Inspirational | 0.952 | Self-assured: Is secure with self, free of doubts | 0.646 |
| 8 | Shows passion for their job | 0.747 | Delegates | 0.952 | Determined: Takes a firm stand, acts with certainty | 0.635 |
| 9 | Able to use intelligence to advantage | 0.546 | Decisive | 0.952 | Diligent: Is persistent, hard working | 0.566 |
| 10 | Shows appropriate rebelliousness | 0.424 | Resilient | 0.833 | Friendly: Shows kindness and warmth | 0.550 |
| 11 |  |  |  |  | Empathic: Understands others, identifies with others | 0.550 |
| 12 |  |  |  |  | Conscientious: Is thorough, organized, and controlled | 0.513 |
| 13 |  |  |  |  | Sensitive: Shows tolerance, is tactful, and sympathetic | 0.412 |
| 14 |  |  |  |  | Outgoing: Talks freely, gets along well with others | 0.312 |
| **Total** |  | **10** |  | **10** |  | **10** |

Participants rated integrity, communication and vision as the three most important traits. Traits were not equally weighted: integrity (1.526) was scored 3.6 times more important than rebelliousness (0.424). No statistically significant gender difference in trait ranking was identified.

**Self-assessment and instrument comparison.** Self-assessment mean scores are shown in Table 2. The distributions of LETS and LTG are assumed to be normal by satisfying both Kolmogorov-Smirnov and Shapiro-Wilk tests, despite the low *n*. Both LETS and LTQ showed Cronbach’s α exceeding 0.7 (See supplement 2), indicating acceptable internal consistency. Limited semantic overlap may occur in the descriptors proposed by subjects in LETS.

**Table 2.**
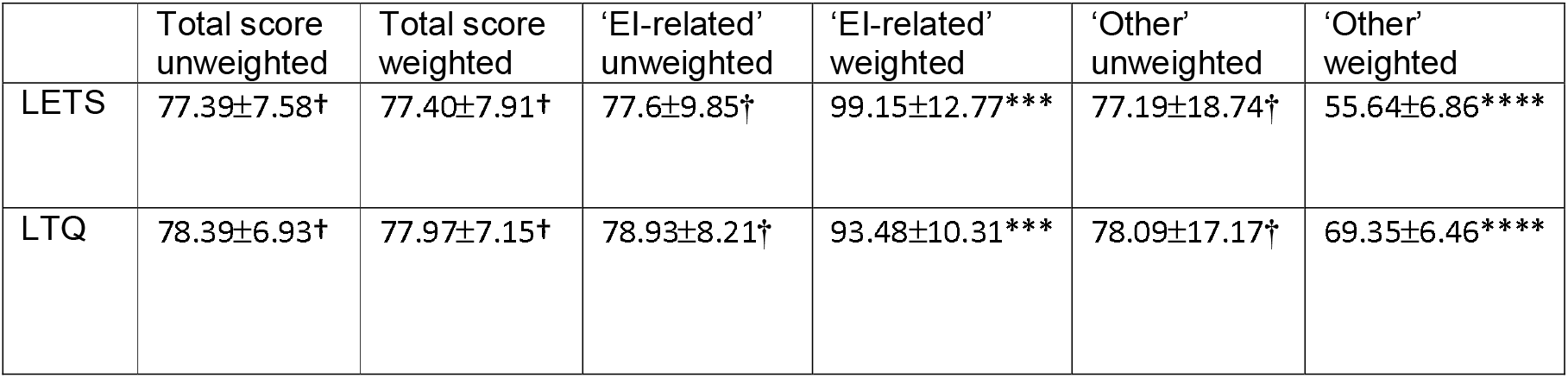
The effect of weighting and separation of EI traits from LETS and LTQ scores (mean. ±**SD) for n=19 subjects. The significance of differences between LETS and LTQ using a paired t-test is: † p=ns, * p<0.05, ** p<0.01, ***p<0.005, ****p<0.001**

|  | Total score unweighted | Total score weighted | 'EI-related' unweighted | 'EI-related' weighted | 'Other' unweighted | 'Other' weighted |
| --- | --- | --- | --- | --- | --- | --- |
| LETS | 77.39±7.58† | 77.40±7.91† | 77.6±9.85† | 99.15±12.77*** | 77.19±18.74† | 55.64±6.86**** |
| LTQ | 78.39±6.93† | 77.97±7.15† | 78.93±8.21† | 93.48±10.31*** | 78.09±17.17† | 69.35±6.46**** |

There was no difference in subject self-scores when total LETS results were compared to total LTG (either raw or weighted scores). This finding supports content validity between LETS (10 items) and LTQ (14 items).

When comparing LETS subset comparisons for raw (unweighted) ‘EI-related’ versus raw (unweighted) ‘other’ scores, there was no significant difference (p=ns, R^2^= 0.0013). However, after LETS scores were weighted for importance as determined by n=19 subjects (Table 2), the subset scores diverged for both ‘EI-related’ and ‘other’ traits (p<0.001, R^2^= 0.9216). LETS and LTG mean scores still correlated when the top five ‘EI-related’ traits were removed and the remaining traits were weighted, demonstrating subset similarity between tools.

### Separation of key ‘EI-related’ traits from ‘other’ traits

Assessment of LETS and LTQ responses (Table 1) indicated a relative preference for EI-related traits, based on Goleman descriptors (9). To determine whether the generic survey (LTQ) differed from the healthcare subjects’ preferred traits (LETS), the top 5 ‘EI-related’ traits were separated from ‘other’ traits (i.e. 50% of 10 traits) (Supplement 3). ‘Other’ traits are not totally exclusive of EI relevance and were deemed not amongst the 5 most ‘EI-related’.

**AI response to leadership suitability**. When asked to rate itself in regard to leadership traits, the comparator replied: “ I don’t have personal experiences or emotions, so I don’t possess leadership traits or self-awareness.” This reply therefore abrogated the possibility of using self-assessment with any of the 3 scales in this study.

## Discussion

### Principal Findings

Senior medical staff identified and ranked ten traits that they considered important qualities of leaders (LETS tool). Neither study participants nor the AI source treated all traits equally. EI-related traits comprised five of the top six preferred traits, and most subjects (and the AI source) did not distinguish EI from its component descriptors. A majority of subjects did not distinguish ‘leadership’ from ‘management’ traits. Characteristics that were considered undesirable in leaders included ‘thirst for power’ and ‘egotistical’.

When using self-scoring to assess comparability of different scales, there was a strong correlation between self-scoring using LETS and LTQ tool mean scores for individuals, suggesting that many generic leadership traits were applicable to healthcare leaders. Trait-weighting did not change the correlation between LETS and LTQ for self-scores, reinforcing the view that many desired leadership traits could be common to both.

Using trait-weighting did not change the correlation between LETS and LTQ, but did reveal differences in mean score for ‘EI-related’ and ‘other’ traits. Of the ten traits nominated by the AI source, five were identical to the LETS top five and reflected EI-related characteristics. These follower-derived priorities are consistent with the social identity theory of leadership, where a previous meta-analysis showed that leaders perceived as prototypical of their group attracted greater trust, endorsement and perceived effectiveness with the strongest effects for ideal-type representations and formal leaders(21). Integrity, communication and empathy may therefore be valued as they signal that a clinical leader represents, and acts for, the professional in-group.

### Comparison with other studies

Since Goleman’s first description of EI there has been considerable interest in determination of the role of an emotional quotient (EQ) in leadership selection and practice (9, 22, 23). Goleman identified five trait domains reflecting desirable qualities in the workplace that were distinctly different to cognitive intelligence (24). In determining the role of EI in healthcare leadership specifically, a study by Bonazza et al found that EI, communication and teamwork were the most important components in a leadership program, with both the junior (medical students) and most senior (faculty members) participants rating EI highest and improving their ratings through structured training (25). The case for development was made earlier by Goleman in a study of leadership theory, who recommended *‘..strengthening these abilities through persistence, practice and feedback from colleagues or coaches’*(22). Goleman made another important observation of relevance to the current work. He found that EI was twice as important as technical skills or IQ. This calculated ratio introduces the concept of relative importance or weighting of characteristics. Calculated ratios now provided by this study found similar ratios for EI-related traits (64% for EI -related and 36% for ‘other’ traits, ratio = 1.78; Table 1). Later work proposed a neurophysiological basis for innate EI was proposed that was dependent on neuronal plasticity and a system of ‘social circuitry’ linked to spindle cell function (26). Although to date unproven, this distinction between ‘innate’ and ‘developed’ EI would justify both selecting leaders with EI and enhancing it through training programs. Enhanced EI may have important implications for the healthcare workforce beyond leadership, by improving morale and reducing burnout (23, 27–29). Although the concept may be desirable, evidence from the literature is inconclusive, suggesting intervention studies on this topic are warranted.

In accord with previous work, the majority of study subjects listed EI as a preferred trait, in addition to listing its components, as did the AI-derived list (14, 20, 25). Further advocacy for EI capability in the healthcare workforce and inclusion in employment selection criteria for leaders is warranted, as is information regarding its definition(30, 31).

Interestingly, the role of power did not arise in this study of healthcare leadership traits (LETS), in Northouse’s LTQ (14), or in AI preferred traits (20). Although, referent power may be attributable to a leader’s social skills (i.e. EI) The five bases of power described by French and Raven serve as a framework to classify components of power used by leaders (32). They include referent (charm), expert (competence), legitimate (status), reward (material benefit) and coercive (penalize) powers. According to this theory, leaders may use any of the five bases alone or in combination to achieve influence, positively or coercively (33) (14).

The present study’s findings also speak to a recent report by Kline, which argued that the dominant NHS employment-relations paradigm, reliant on policies, procedures and training, has been largely ineffective, and called instead for accountability, inclusive leadership and the debiasing of systems(4). In reference to the current study, the attributes followers valued most (integrity, communication, emotional intelligence and empathy) provide concrete content to the kind of trustworthy, inclusive leadership Kline links to psychological safety and a healthier culture.

### Strengths and Limitations

The principal strength of the present study is a follower-driven design: leadership attributes were generated by senior medical staff who will be led by external frameworks. The study deliberately triangulated across three independent sources (qualitative focus groups (LETS), established published instrument (LTQ) and a large language model (LAIT), yielding a highly-consistent, top-five profile and supported convergent validity. The use of both raw and empirically-weighted scoring is uncommon in leadership-trait literature and exposed content validity differences that a single-study method may not recognise.

Several limitations apply. It is uncertain whether the study findings are representative of all senior medical staff, given limited power of the study at 45% for α=0.05. Findings should be treated as hypothesis-generating rather than confirmatory. Recruitment was opportunistic and single-site, drawn only from medical staff from a Melbourne-based hospital, which may limit its generalisability beyond comparable populations and tertiary settings uncertain. Self-scoring against LETS and LTQ may be subject to social-desirability bias. Finally, the comparison of the LETS with LAIT (AI comparator) reflects a single query to one model (GPT-3.5) and may change with the evolution of AI.

### Implications

This finding has implications for the selection of leaders, in that simple binary scoring of characteristics may overscore those with less important traits and underscore those with desired traits. In progressively complex healthcare systems, effective leadership is with the need for capable healthcare leaders, a recent scoping review argues that, in increasingly conceptualised as a set of broad goals (i.e. collaboration, capacity-building, and continuous innovation), rather than as a fixed competency checklist(34). The follower-weighted LETS approach aligns with this perspective by recognising that different leadership traits contribute unequally to perceived effectiveness.

Practically, the LETS scale offers a simple tool for: (1) supporting weighted, follow-informed selection criteria for clinical leadership appointments, (2) structuring 360 degree feedback and leadership-development programmes around an EI-related cluster that emerged as dominant in both human and AI-derived trait sets and (3) self-reflective review of performance by incumbent leaders.

### Future Research

This study requires validation in larger, multi-site cohorts, potentially including nursing, allied-health, junior medical staff, with enhanced statistical power. The LAIT (AI-comparator) requires investigation to determine validation with newer and various models, given that AI leadership “capabilities” lack experiential and emotional grounding. We also propose a longitudinal study to determine the predictive validity of LETS-based selection on downstream outcomes such as staff engagement, psychological safety and patient experience.

## Conclusion

Healthcare professionals have a clear preference for emotionally-intelligent leaders. A selection tool such as LETS offers senior medical staff some reassurance that leaders can be chosen to meet their needs, thereby reinforcing the leader-follower relationship. Furthermore, the LETS tool may also be used to both assess acceptability of incumbent leaders and assist with self-appraisal of those aspiring to leadership roles. Given the present study’s exploratory scale, these conclusions warrant confirmation in larger, more diverse healthcare populations.

## Statements

## Data Availability

All data produced in the present study are available upon reasonable request to the authors

## Acknowledgements

We thank the senior medical staff who participated in the focus groups and survey. We thank Prof Alison Wolf (KCL) for supervision of the project.

## Use of AI

OpenAI ChatGPT (GPT-3.5) was used on 22 July 2023 as a comparator data source only (the LAIT trait list; Supplement 1). No generative AI was used in the writing, editing, or analysis of this manuscript. The authors take full responsibility for all content.

## Contributors

John Wilson conceived and designed the study, conducted the focus groups and survey, performed the analysis, drafted the manuscript, and is guarantor. Angela Michaelis contributed to the critical revision and drafting of the manuscript for important intellectual content. Mary-Ellen Miller contributed to the study concept, interpretation and discussion of the findings.

## Funding

This research received no specific grant from any funding agency in the public, commercial, or not-for-profit sectors.

## Competing interests

None declared.

## Ethics approval

The study was approved by the Monash University Human Research Ethics Committee (project 35811, approved July 2023) and was conducted in accordance with the Declaration of Helsinki. All participants gave informed consent.

## Data availability statement

De-identified survey data and the LETS/LTQ scoring spreadsheet are available from the corresponding author on reasonable request. Raw focus-group transcripts are not shared, to protect the identity of participants given the small and potentially identifiable cohort. The full AI prompt and response are provided in Supplement 1.

## Patient consent for publication

Not applicable.

## Supplement 1

***Explain in 100 words what the risk is for leadership and decision-makers posed by A.I.***

***(22 July 2023, 4.20pm)***

AI poses significant risks for leadership and decision-makers. Firstly, reliance on AI may lead to a loss of human judgment and critical thinking, diminishing leaders’ ability to make independent, ethical decisions. Secondly, biased algorithms can perpetuate discrimination and inequality, causing reputational damage and legal consequences. Thirdly, AI’s complexity may obscure decision processes, making accountability challenging. Lastly, AI’s rapid evolution demands leaders stay updated to leverage its potential fully, while unpreparedness risks being outpaced by competitors. To navigate these risks successfully, leaders must exercise caution, ensure transparency, embrace AI responsibly, and combine human expertise with AI insights.

***What are the best 10 traits of leaders? 22 July 2023, 4.27pm***

1. **Visionary**: Great leaders have a clear vision of what they want to achieve and can articulate it to their team. They inspire others by painting a compelling picture of the future and motivating everyone to work towards that shared vision.
2. **Integrity**: Leaders with integrity earn the trust and respect of their team members. They demonstrate strong moral and ethical principles, and their actions align with their words. Trust is the foundation of a strong team, and integrity helps to build that trust.
3. **Decisiveness**: Making tough decisions is a crucial aspect of leadership. Effective leaders are capable of analyzing information, considering alternatives, and making timely decisions. They understand that indecisiveness can lead to stagnation and uncertainty within the team.
4. **Empathy**: Being able to understand and relate to the feelings and perspectives of others is a hallmark of a good leader. Empathy allows leaders to connect with their team members on a deeper level, fostering a supportive and compassionate work environment.
5. **Effective Communication**: Leaders must be excellent communicators. They can convey their ideas clearly and actively listen to others. Strong communication skills help leaders resolve conflicts, provide feedback, and keep the team aligned with organizational goals.
6. **Confidence**: Confidence inspires confidence in others. A self-assured leader instills a sense of belief and optimism in their team, even during challenging times. It helps create a positive atmosphere where team members feel more secure and motivated.
7. **Adaptability**: The ability to adapt to changing circumstances and embrace new ideas is crucial for leaders. Markets, industries, and teams evolve, and leaders who can adjust their strategies and approach will stay relevant and effective.
8. **Accountability**: Great leaders take responsibility for their actions and the outcomes of their decisions. They hold themselves and their team accountable for their performance, learning from failures, and celebrating successes.
9. **Resilience**: Leaders face numerous obstacles and setbacks, but their resilience helps them bounce back and persevere through challenging times. Resilient leaders inspire their teams to keep pushing forward despite difficulties.
10. **Empowerment**: Effective leaders empower their team members, providing them with autonomy and opportunities for growth. They delegate tasks and responsibilities, recognizing that fostering a sense of ownership in team members boosts motivation and productivity.

Remember that leadership is a continuous learning process, and while possessing these traits is important, being open to growth and development is equally essential for becoming a successful leader.

***For these 10 leadership qualities, what weighting for importance do each of these have?***

***22 July 2023, 4.37pm***

Assigning precise weightings to the importance of leadership qualities can be subjective and may vary depending on the specific context and organization. However, here’s a general guideline that reflects the relative significance of each trait:

1. **Visionary: 9/10** - Having a clear and inspiring vision is critical for guiding the team towards a shared goal. Without a strong vision, it’s challenging to align efforts effectively.
2. **Decisive: 8/10** - Timely decision-making is essential for maintaining momentum and avoiding stagnation. Decisiveness allows leaders to take advantage of opportunities and address challenges promptly.
3. **Empathetic: 9/10** - Empathy is crucial for building strong relationships and fostering a positive, supportive team culture. It boosts morale and encourages collaboration.
4. **Effective Communicator: 9/10** - Communication is the foundation of successful leadership. Clear and open communication ensures that everyone understands expectations and can contribute effectively.
5. **Integrity: 9/10** - Trust is the cornerstone of leadership, and integrity is the bedrock of trust. Without integrity, a leader’s influence will erode over time.
6. **Resilient: 7/10** - While resilience is valuable, it may not always be the most critical factor. However, in challenging environments, a leader’s ability to bounce back and motivate others is vital.
7. **Inspirational: 8/10** - Inspirational leaders can motivate teams to go above and beyond. Cultivating inspiration drives creativity and dedication.
8. **Delegation: 8/10** - Effective delegation empowers team members, promotes growth, and prevents burnout. Leaders who fail to delegate risk becoming overwhelmed and stifling their team’s potential.
9. **Problem Solvers: 8/10** - Problem-solving skills are essential for overcoming obstacles and driving innovation. Leaders who excel in this area can navigate complex situations effectively.
10. **Emotional Intelligence: 9/10** - Emotional intelligence is crucial for understanding and managing relationships. It greatly impacts how leaders handle conflicts and build strong teams.

Remember, the relative importance of each trait can change based on factors like the nature of the organization, the team’s composition, and the challenges faced. Great leaders often possess a blend of these qualities and know when to emphasize certain traits based on the situation at hand. Leadership is a dynamic journey that requires constant self-awareness, learning, and adaptation.

***Please rate yourself out of 10 for each of these leadership traits 22 July 2023, 4.40pm***

As an AI language model, I don’t have personal experiences or emotions, so I don’t possess leadership traits or self-awareness. However, I have been trained on a vast dataset that includes information about leadership qualities, and I can provide information and insights on these traits based on that training.

If you have any questions or need further information on leadership traits or any other topic, feel free to ask!

***How do good leaders best use AI? 22 July 2023, 5.09pm***

Good leaders can leverage AI in various ways to enhance their leadership and improve organizational outcomes. Here are some ways in which good leaders can best use AI:

1. **Data-Driven Decision Making:** AI can analyze vast amounts of data quickly and accurately. Good leaders can use AI-powered analytics to make more informed and data-driven decisions, leading to better strategies and outcomes.
2. **Personalized Customer Experiences:** AI can help leaders understand customer preferences and behavior better. By using AI in marketing and customer service, leaders can deliver personalized experiences, enhancing customer satisfaction and loyalty.
3. **Process Automation:** AI can automate repetitive and mundane tasks, freeing up time for leaders and their teams to focus on more strategic and value-added activities. This increases overall efficiency and productivity.
4. **Talent Management:** AI can assist leaders in identifying top talent, predicting employee attrition, and analyzing workforce data. This enables leaders to make better decisions related to talent recruitment, development, and retention.
5. **Predictive Maintenance:** In industries with physical assets, AI-powered predictive maintenance can help leaders optimize maintenance schedules and reduce downtime, leading to cost savings and improved operational efficiency.
6. **Market and Competitive Analysis:** AI can analyze market trends, competitors, and consumer behavior to provide leaders with valuable insights. This enables leaders to make more informed decisions about market positioning and potential opportunities.
7. **Innovation and R&D:** AI can aid in the discovery of patterns and trends in research and development efforts. It can also assist in identifying potential breakthroughs, leading to more efficient innovation processes.
8. **Risk Management:** AI can help leaders in risk assessment and management by identifying potential risks and vulnerabilities. This empowers leaders to proactively address challenges and mitigate potential threats.
9. **Customer Feedback Analysis:** AI-powered sentiment analysis can help leaders gain a deeper understanding of customer feedback and preferences. This information can be used to improve products, services, and overall customer satisfaction.
10. **Employee Engagement and Well-being:** AI can be used to monitor employee engagement and well-being by analyzing data related to work patterns and stress levels. Leaders can use this data to create a healthier and more supportive work environment.

However, while AI offers numerous benefits, good leaders should also be mindful of potential challenges, including data privacy concerns, ethical implications, and the need for proper governance. They should approach AI implementation with a strategic and responsible mindset, ensuring that it aligns with the organization’s values and goals. Additionally, good leaders should actively foster a culture of learning and adaptability to ensure that their teams can embrace and leverage AI effectively.

***Does ChatGPT use information on my computer to formulate a reply to my questions?***

***23 July 2023, 11.19am***

No, ChatGPT does not have access to any information on your computer or any external data source to formulate replies to your questions. As an AI language model, my responses are generated based solely on the vast amount of data I have been trained on up until my knowledge cutoff date in September 2021.

When you interact with ChatGPT, your questions are sent to the OpenAI servers, where the model processes the input and generates a response. The responses are generated on-the-fly and are not influenced by any specific information about you or your device.

It’s important to note that while ChatGPT doesn’t have memory of past interactions within a session, OpenAI may collect and retain data as described in its data usage policy for various purposes, including improving the system and ensuring the quality of its services. However, any data that is collected is handled with privacy and security measures in place. If you have concerns about data privacy, I recommend reviewing OpenAI’s data usage policy for more information.

Supplement 2.

***Internal consistency using Cronbach’s α (n=16 subjects)***

(35, 36)

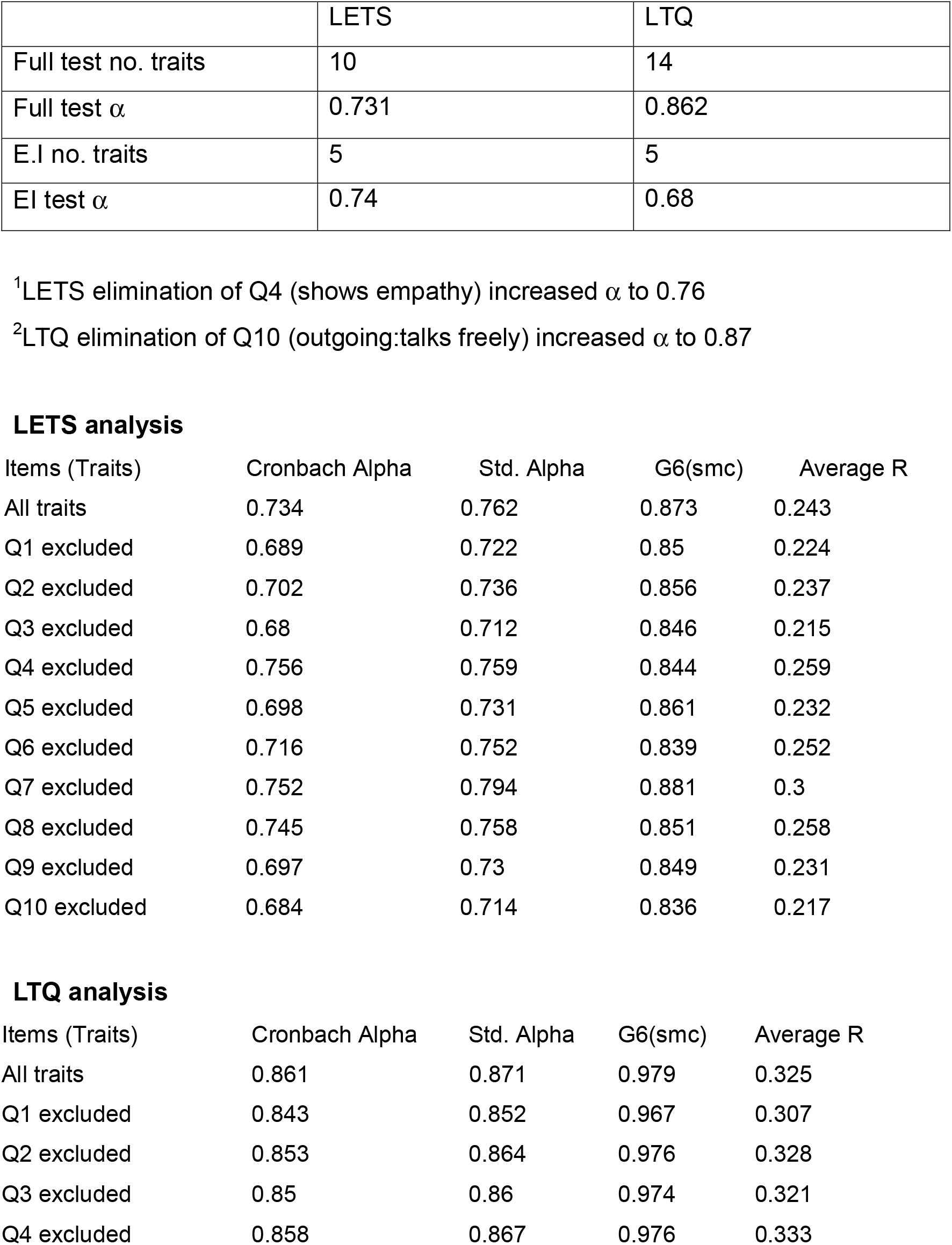
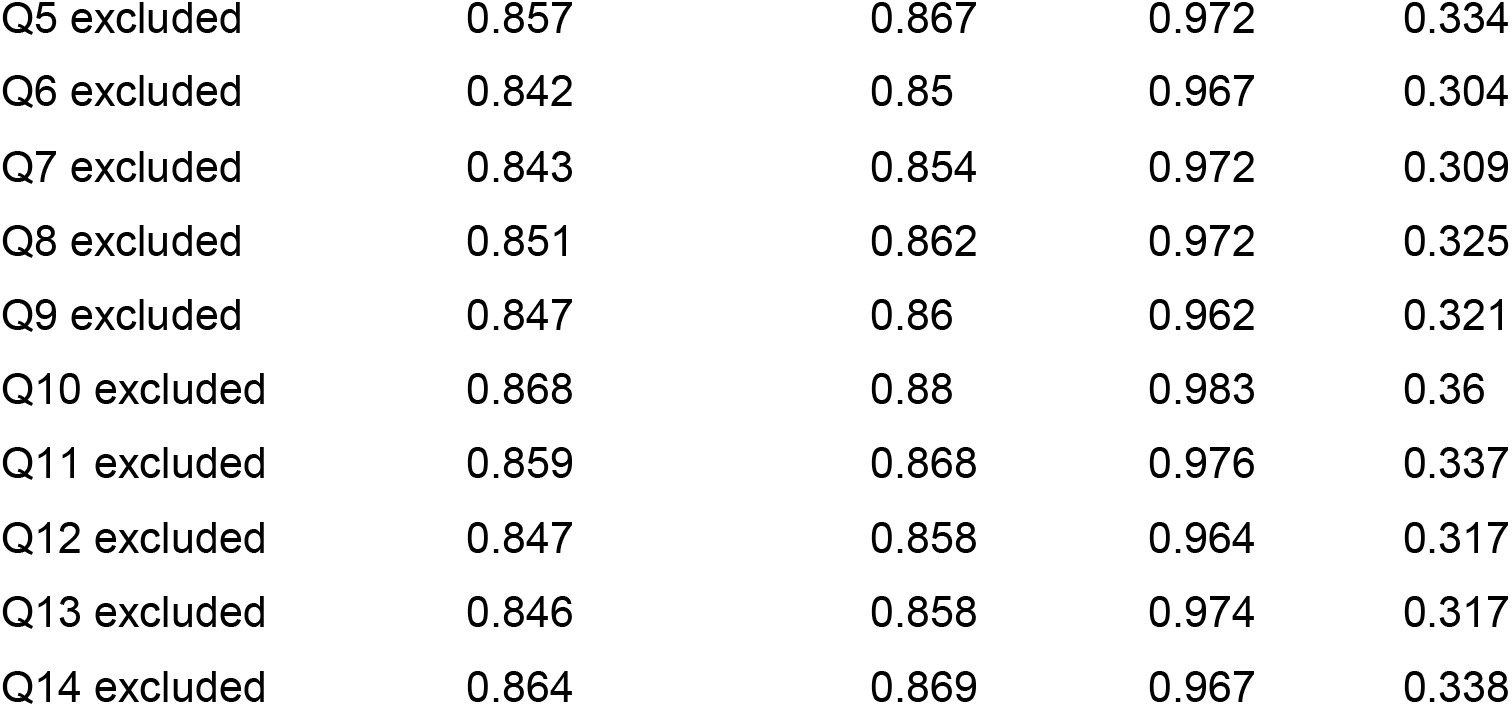

Supplement 3.

**Table 3.**
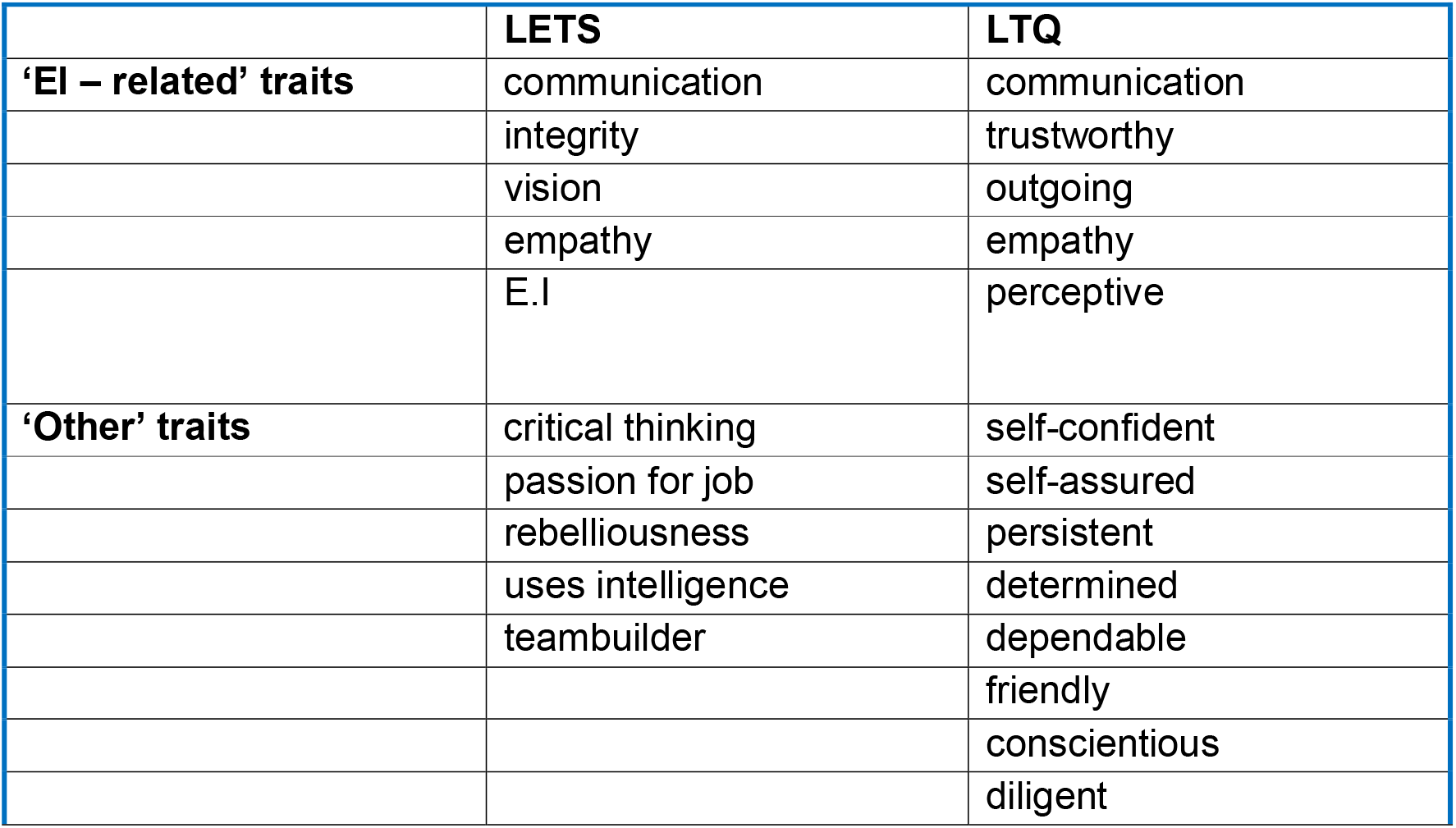
Arbitrary separation of top 5 ‘EI-related’ traits from ‘other’ traits for LETS and LTQ.

## Notes

### Competing Interest Statement

The authors have declared no competing interest.

### Author Declarations

The study was approved by the Monash University Human Ethics Research Committee (Project 35811, approved July 2023)

